# What is known from the existing literature about the available treatments for pelvic floor dysfunction among female athletes? A scoping review protocol

**DOI:** 10.1101/2021.05.10.21256950

**Authors:** Silvia Giagio, Stefano Salvioli, Paolo Pillastrini, Marika Fiorucci, Tiziano Innocenti

## Abstract

**Background:** Pelvic floor dysfunction (PFD) is a term used to describe a variety of symptoms, signs and conditions involving different impairments on the pelvic floor muscles.

The existing literature suggests that some sports may lead to a higher risk of developing PFD, in particular among female athletes. Despite a recent scoping review highlighted a great number of studies dealing with epidemiologic data on this topic, no study has been conducted to map the available treatments.

In this framework, the aim of the present scoping review will be to map and summarize the literature to identify the available evidence concerning the treatments for PFD among female athletes.

**Inclusion criteria:** Studies considering female athletes practicing sports at any performance level with any type of PFD will be eligible for inclusion. Any treatment options (i.e. preventive, conservative, surgery) reported by each study and any context will be considered.

**Methods:** This scoping review will be performed in accordance with the Joanna Briggs Institute methodology. MEDLINE, Cochrane Central, Scopus, CINAHLComplete, Embase, PEDro and SPORTDiscus database will be searched from inception to May 2021. Additional records will be identified through searching in grey literature and the reference lists of all relevant studies. No study design, publication type, data and language restrictions will be applied.

Two reviewers will independently screen all abstracts and full-text studies for inclusion. A data collection form will be developed by the research team to extract the characteristics of the studies included. A tabular and accompanying narrative summary of the information will be provided.

**Conclusions:** This will be the first scoping review to provide a comprehensive overview of the topic. The results will add meaningful information for clinicians in the management of PFD among female athletes. Furthermore, any knowledge gaps of the topic will be identified. The results of this research will be published in a peer-reviewed journal and will be presented at relevant (inter)national scientific events.

## INTRODUCTION

Pelvic floor dysfunction (PFD) is a term used to describe symptoms, signs, and conditions primarily affecting women, with or without moderate-to-severe impairment of the pelvic floor muscles^1^.

Among the athlete population practicing different sports, a recent scoping review^2^ highlighted a wide range of published studies regarding the epidemiologic data of PFD. In particular, the majority of the included studies (n=83, 83%) focused only on the females.

Despite these findings, the increasing interest on the topic^2^ and the high prevalence of dysfunctions that emerged from several reviews^3–5^, there is little research regarding the management of PFD within this group.

What are the available evidence-based treatments for female athletes with PFD?

To the authors’ knowledge, no study has been conducted in this regard and as a consequence, there is no a comprehensive overview both for clinicians and researchers.

In this context, the aim of the present study will be to summarize the literature to identify the available evidence of treatment options for PFD in female athletes. This clinical data synthesis could add meaningful information for the overall management of the athlete.

As maintained by the Joanna Briggs Institute (JBI)^6^, scoping review approach may be used to map and clarify key concepts, identify gaps in the research knowledge base, and report on the types of evidence that address and inform practice in the field. These aims corresponded to the objectives of this project. For this reason, other types of review, such as systematic reviews, umbrella reviews or rapid reviews, were not deemed methodologically effective.

## METHODS

The present scoping review will be conducted in accordance with the JBI methodology^6^ for scoping reviews.

The Preferred Reporting Items for Systematic reviews and Meta-Analyses extension for Scoping Reviews (PRISMA-ScR)^7^ Checklist for reporting will be used.

### Research team

To facilitate robust and clinically relevant review findings, the research team will include individuals with expertise in evidence synthesis, quantitative and qualitative research methodology, sport physiotherapy and pelvic floor rehabilitation.

### Review question(s)

We formulated the following research question: “What is known from the existing literature about the treatments for PFD among female athletes?”

In particular, the objectives of this scoping review will be to:

1. Provide a comprehensive overview of all studies dealing with PFD treatments for the female athletes;
2. Identify and summarize studies according to the type of sport, PFD classified on the basis of the International Continence Society (ICS) standardized terminology, type of treatment and eventually other subgroups that could emerge from the analysis of the included studies.
3. Identify any knowledge gaps of the topic.

### Inclusion criteria

Studies will be eligible for inclusion if they meet the following Population, Concept, and Context (PCC) criteria.

▪ *Population*. We will include female athletes of any age, practicing any type of sport and performance level (e.g., professional/elite, amateurs/master/recreational athletes) with any type of PFD. The definition of “athlete” used in an individual study as the main criterion will be considered.
▪ *Concept*. Any treatment options (i.e. preventive, conservative, surgery) reported by each study will be considered.
▪ *Context*. This review will consider studies conducted in any context.

*Sources*. This scoping review will consider any study designs or publication type. No time, geographical, setting and language restrictions will be applied.

### Exclusion criteria

Studies that do not meet the specific PCC criteria will be excluded.

### Search strategy

An initial limited search of MEDLINE was undertaken to identify articles on the topic and then index terms used to describe the articles was used to develop a full search strategy for MEDLINE (see **Table 1**, which displays the search strategy for MEDLINE database). The search strategy, including all identified keywords and index terms will be adapted for use in Cochrane Central, Scopus, CINAHLComplete, Embase, PEDro and SPORTDiscus.

**Table 1.** Search strategy for MEDLINE.

| **MEDLINE** |  |
| --- | --- |
| Search conducted on | 9<sup>th</sup> May 2021 |
| Results | 967 |
| Search strategy | (("pelvic floor disorders"[MeSH Terms] OR ("Pelvic"[All Fields] AND "Floor"[All Fields] AND "disorders"[All Fields]) OR "pelvic floor disorders"[All Fields] OR ("pelvic floor"[MeSH Terms] OR ("Pelvic"[All Fields] AND "Floor"[All Fields]) OR "pelvic floor"[All Fields]) OR ("urinary incontinence"[MeSH Terms] OR ("Urinary"[All Fields] AND "Incontinence"[All Fields]) OR "urinary incontinence"[All Fields]) OR ("Dyspareunia"[MeSH Terms] OR "Dyspareunia"[All Fields]) OR ("faecal incontinence"[All Fields] OR "fecal incontinence"[MeSH Terms] OR ("Fecal"[All Fields] AND "Incontinence"[All Fields]) OR "fecal incontinence"[All Fields]) OR ("intestinal obstruction"[MeSH Terms] OR ("Intestinal"[All Fields] AND "Obstruction"[All Fields]) OR "intestinal obstruction"[All Fields]) OR ("pelvic organ prolapse"[MeSH Terms] OR ("Pelvic"[All Fields] AND "Organ"[All Fields] AND "Prolapse"[All Fields]) OR "pelvic organ prolapse"[All Fields]) OR ("Cystocele"[MeSH Terms] OR "Cystocele"[All Fields] OR "cystoceles"[All Fields] OR "cystocoele"[All Fields] OR "cystocoeles"[All Fields]) OR ("rectal prolapse"[MeSH Terms] OR ("Rectal"[All Fields] AND "Prolapse"[All Fields]) OR "rectal prolapse"[All Fields]) OR ("uterine prolapse"[MeSH Terms] OR ("Uterine"[All Fields] AND "Prolapse"[All Fields]) OR "uterine prolapse"[All Fields]) OR ("visceral prolapse"[MeSH Terms] OR ("Visceral"[All Fields] AND "Prolapse"[All Fields]) OR "visceral prolapse"[All Fields]) OR ("Vaginismus"[MeSH Terms] OR "Vaginismus"[All Fields]) OR "pelvic pain"[MeSH Terms] OR ("pelvic floor disorde*[Title/Abstract] OR ((("pelvic floor"[MeSH Terms] OR ("Pelvic"[All Fields] AND "Floor"[All Fields]) OR "pelvic floor"[All Fields]) AND "desea*[Title/Abstract]) OR "pelvic floor dysfunction"[Title/Abstract] OR "pelvic floor muscle*[Title/Abstract] OR "pelvic diaphrag*[Title/Abstract] OR "urinary incontinence"[Title/Abstract] OR "urinary urge incontinence"[Title/Abstract] OR "urinary stress incontinence"[Title/Abstract] OR ((("urinary tract"[MeSH Terms] OR ("Urinary"[All Fields] AND "tract"[All Fields]) OR "urinary tract"[All Fields] OR "Urinary"[All Fields]) AND "reflex incontinence"[Title/Abstract]) OR "Dyspareunia"[Title/Abstract] OR "fecal incontinence"[Title/Abstract] OR "bowel incontinence"[Title/Abstract] OR "anus prolapse"[Title/Abstract] OR "intestinal obstruction"[Title/Abstract] OR "pelvic organ prolapse"[Title/Abstract] OR "Cystocele"[Title/Abstract] OR "rectal prolapse"[Title/Abstract] OR "uterine prolapse"[Title/Abstract] OR "visceral prolapse"[Title/Abstract] OR "vaginal prolapse"[Title/Abstract] OR "Vaginismus"[Title/Abstract] OR "pelvic pain"[Title/Abstract]) OR "anal incontinence"[Title/Abstract]) AND ("sport s"[All Fields] OR "sports"[MeSH Terms] OR "sports"[All Fields] OR "sport"[All Fields] OR "sporting"[All Fields] OR "physical fitness"[MeSH Terms] OR ("spor*[Title/Abstract] OR "athlet*[Title/Abstract] OR "physical fitness"[Title/Abstract] OR "Fitness"[Title/Abstract])) AND ("womans"[All Fields] OR "Women"[MeSH Terms] OR "Women"[All Fields] OR "Woman"[All Fields] OR "women s"[All Fields] OR "womens"[All Fields] OR "Female"[MeSH Terms] OR "Woman"[Title/Abstract] OR "Women"[Title/Abstract] OR "Female"[Title/Abstract]) |

In addition, also grey literature (e.g. Google scholar, direct contact with experts in the field of PFD and sports medicine) and the reference lists of all relevant studies will be searched.

### Study selection

Once the search strategy has been successfully completed, search results will be collated and imported to EndNote V.X9 (Clarivate Analytics, PA, USA). Duplicates will be removed using the EndNote deduplicator before the file containing a set of unique records is made available to reviewers for further processing (i.e. study screening and selection). The review process will consist of two levels of screening using Rayyan QCRI online software^8^: (1) a title and abstract review and (2) a full-text screening. For both levels, two authors independently screened the articles with conflicts resolved by a third author.

Reasons for the exclusion of any full-text source of evidence will be recorded and reported in the scoping review report. The results of the search will be reported in full in the final scoping review and presented in the latest published version of the Preferred Reporting Items for Systematic Reviews and Meta-analyses (PRISMA) flow diagram^9^.

### Data extraction

An ad-hoc data extraction form will be developed by the reviewers and key information (e.g. authors, country, year of publication, PFD, sport, type of treatment) on the selected articles will be collected. A draft extraction tool is provided (see **Table 2**, which illustrates the data extraction instrument).

**Table 2.** Data extraction instrument: draft.

| **Study details** |  |
| --- | --- |
| Author(s) |  |
| Year of publication |  |
| Study design |  |
| **Population** |  |
| Sample size |  |
| Age |  |
| “Athlete” definition described by authors | e.g. professional/elite, amateurs/master/recreational athletes) |
| Pelvic floor dysfunction |  |
| Sport |  |
| **Concept** |  |
| Treatment | (i.e. preventive, conservative, surgery) |
| **Context** |  |
| Country |  |

This form will be reviewed by the research team and pre-tested by all reviewers before implementation to ensure that the form captures the information accurately. Charting results is commonly an iterative process during scoping reviews; other data can be added to this form according to the subgroups that could emerge from the analysis of the studies included. Modifications will be detailed in the full scoping review.

### Data synthesis

The results will be presented in two ways:

1. Numerically. Studies identified and included will be reported, and the description of the search decision process will be mapped. In addition, extracted data will be summarized in tabular and diagrammatic form. Charting the results is based on an iterative approach, and further categories may be added if considered appropriate.
2. Thematically. A thematic summary will be performed pertaining to themes and key concepts relevant to the research questions and according to subgroups (e.g. PFD, sport, performance level, type of treatment) and to others that could emerge.

## Data Availability

All data are included in the report.

## Acknowledgements

Not applicable.

## List of abbreviations

ICS: International Continence Society
JBI: Joanna Briggs Institute
PCC: Population-concept-context
PFD: Pelvic floor dysfunction
PRISMA: Preferred Reporting Items for Systematic Reviews and Meta-analyses
PRISMA-ScR: The Preferred Reporting Items for Systematic reviews and Meta-Analyses extension for Scoping Reviews

